# Characterizing the Stability of Radiomics-Derived Tumor Habitats Using Image Perturbation in Head and Neck Cancer

**DOI:** 10.64898/2026.05.30.26354532

**Authors:** Oya Altinok, Asim Waqas, Ghulam Rasool, Matthew B. Schabath, Albert Guvenis

## Abstract

Tumor habitat imaging aims to capture intratumoral heterogeneity by grouping voxels with similar radiomic properties into spatially coherent subregions. However, radiomic features are known to be sensitive to small variations in image acquisition and processing, which can affect the stability of the resulting habitat maps. Feature repeatability is usually evaluated using test–retest scans, but such data are rarely available in clinical practice. To overcome this, we adopted an image perturbation framework, which simulates test–retest conditions by applying small, controlled changes to a single image. In head and neck cancer (HNC), where imaging is further complicated by complex anatomy, dental artifacts, and variability in tumor delineation, dedicated stability analyses are still missing. In this study, we evaluated how the repeatability of radiomic features affects habitat stability in 390 oropharyngeal cancer patients (discovery cohort). For each patient, 11 perturbed CT volumes were generated using small in-plane rotations, sub-voxel translations, and tumor-adaptive Gaussian noise. Ninety-three radiomic features were extracted from each image set, and their repeatability was assessed using the lower confidence limit of the intraclass correlation coefficient (ICC_LCL), grouped into poor, moderate, good, and excellent categories. Tumor habitats were then generated using K-means clustering (*H* = 3) for each feature subset, and habitat stability was measured by the Dice similarity coefficient (DSC) between habitat maps obtained from original and perturbed images. Overall, 48.4% of features were poorly repeatable and only 6.5% reached the excellent category, with first-order features being more stable than texture-based ones. Habitat stability followed a clear monotonic trend with feature repeatability: the median DSC was 0.93 for habitats generated from excellent features, 0.84 for good features, 0.75 for moderate features, and dropped to 0.41 for poorly repeatable features. Habitats generated using all features (without any repeatability-based filtering) yielded an intermediate median DSC of 0.52. All pairwise comparisons between feature subsets were statistically significant (*p* < 0.001). To evaluate the generalizability of these findings, the analysis was repeated in an independent external validation cohort of 372 oropharyngeal cancer patients treated at the H. Lee Moffitt Cancer Center. The stability classification showed substantial feature-level concordance between the discovery and validation cohorts (overall agreement 67.7%, quadratic-weighted Cohen’s kappa = 0.78), with no feature shifting by more than two stability classes. The habitat-stability hierarchy was fully preserved in the validation cohort (median DSC of 0.87, 0.73, 0.69, and 0.39 for excellent, good, moderate, and poor features, respectively; all pairwise *p* < 0.001). These results show that selecting features with higher repeatability clearly improves the spatial consistency of habitat maps in HNC, and support the use of perturbation-based stability analysis as a routine step in habitat imaging studies.

## 1 Introduction

Solid tumors are rarely uniform. Different regions within the same tumor often show distinct biological behavior, including differences in cell density, blood supply, oxygen levels, and necrosis [1, 2, 3]. This intratumoral heterogeneity is a known driver of treatment resistance and is linked to worse clinical outcomes [4, 5, 6, 7]. Capturing this variation noninvasively is therefore an important goal in oncology, and medical imaging offers a practical way to do so on a whole-tumor scale.

One imaging-based approach to study heterogeneity is habitat analysis, which divides a tumor into subregions sharing similar imaging properties [8, 9]. Habitats are typically built by computing radiomic features at the voxel level and then grouping voxels with similar feature values through unsupervised clustering. This approach has been applied to several cancer types and clinical tasks, including outcome prediction in ovarian cancer, malignancy assessment in lung lesions, and biopsy guidance [10, 11, 12]. Radiotherapy planning, where treatment-resistant subregions could potentially receive higher doses, has also been explored as a possible application [13].

A key limitation of habitat imaging is that radiomic features are sensitive to many factors beyond the underlying tumor biology, such as scanner type, reconstruction settings, voxel size, and segmentation [14, 15, 16]. Because habitats are derived directly from these features, any instability at the feature level can affect the resulting habitat maps. Repeatability is usually evaluated using test–retest scans, but such datasets are rarely available in clinical practice [17]. To work around this, image perturbation has been proposed as an alternative. By applying small, plausible changes to a single image, such as rotation, translation, or noise addition, perturbation can mimic the differences expected between repeated scans [18]. Earlier work has shown that perturbation chains produce feature-stability rankings comparable to those from real test–retest data, and that selecting only repeatable features leads to more stable habitat maps in lung and liver tumors [18, 19, 20]. Findings from these studies also suggest that the set of stable features depends on the anatomical site, meaning that results obtained for one tumor type cannot be directly transferred to another.

Head and neck cancer (HNC) presents specific imaging challenges, including complex anatomy, dental artifacts, and notable variability in tumor delineation [21], all of which may influence the stability of voxel-wise radiomic features. Despite this, dedicated repeatability and habitat-stability analyses for HNC are still missing in the literature. The aim of this study is to address this gap. First, we use an image perturbation framework to assess the repeatability of voxel-wise radiomic features in HNC. Second, we examine how feature-level repeatability translates into habitat-level stability by comparing habitat maps obtained from original and perturbed images, using the Dice similarity coefficient as the main metric. Third, we externally validate both the proposed stability classification and the resulting habitat-stability hierarchy in an independent institutional cohort of oropharyngeal cancer patients. By doing so, we aim to identify which features can be reliably used for habitat construction in HNC and to provide a basis for more robust habitat-based biomarkers in this cancer type.

## 2 Materials and Methods

This study used two independent OPC cohorts to evaluate the effect of voxel-wise radiomic feature repeatability on habitat stability. The discovery cohort was used to define feature repeatability classes, whereas the external validation cohort was used to test the generalizability of these classes and their effect on downstream habitat stability. The overall workflow of this study is shown in Figure 1.

**Figure 1:**
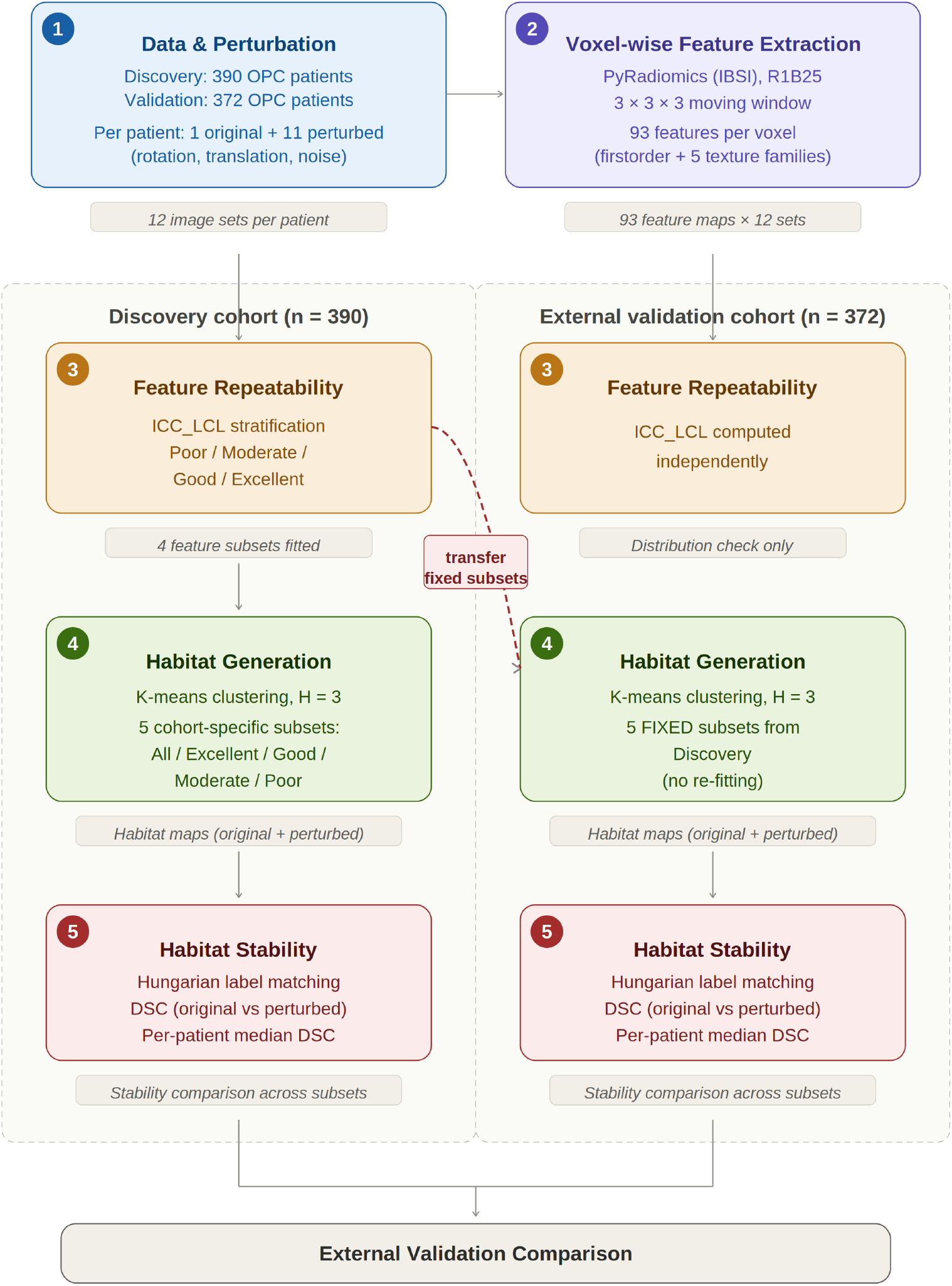
Overall workflow of the study. Steps 1–2 (data acquisition, image perturbation, and voxel-wise feature extraction with PyRadiomics) are applied identically to both cohorts. The pipeline then splits into two parallel arms: in the discovery cohort (*n* = 390), feature repeatability is assessed using ICC_LCL stratification (Step 3) and feature subsets are fitted; in the external validation cohort (*n* = 372), habitat generation (Step 4) uses the fixed feature subsets transferred from the discovery cohort without re-fitting. Habitat stability (Step 5) is quantified in both cohorts using the Dice similarity coefficient (DSC). The two arms converge in the external validation comparison.

### 2.1 Study Cohorts

Two independent cohorts originating from different institutions were used in this study. The discovery cohort consisted of 390 patients diagnosed with oropharyngeal cancer (OPC) and was obtained from The Cancer Imaging Archive (TCIA) [22, 23, 24]. The dataset included pre-treatment contrast-enhanced CT images with expert segmentation labels. The gross primary tumor volume (GTVp) was used as the region of interest. This cohort was used to establish the stability classification of voxel-wise radiomic features and the corresponding habitat-stability hierarchy.

The external validation cohort consisted of 372 OPC patients treated at H. Lee Moffitt Cancer Center & Research Institute and scanned with an independent set of CT acquisition protocols. This study was approved by an Institutional Review Board (Advarra Inc.). For this cohort, intratumoral segmentation was generated by an imaging scientist with 14 years of experience using a single-click ensemble segmentation approach implemented in HealthMyne software [25].

The same image perturbation, feature extraction, habitat generation, and stability quantification pipelines were applied to both cohorts. The stability classification derived from the discovery cohort was treated as the reference and applied to the validation cohort without refitting, in order to assess its external generalizability under three complementary perspectives, as detailed in Section 2.7.

### 2.2 Image Preprocessing and Perturbation Framework

To assess radiomic feature repeatability in the absence of test–retest imaging data, a perturbation-based framework was implemented. Perturbation methods simulate test–retest conditions by introducing controlled variations to the original images, enabling repeated feature extraction under varying conditions. This approach has been standardized by Zwanenburg et al. [26] and is widely used in radiomics for evaluating feature robustness. It provides a reproducible strategy for evaluating how radiomic features respond to imaging variability. The perturbation framework is shown in Figure 2.

**Figure 2:**
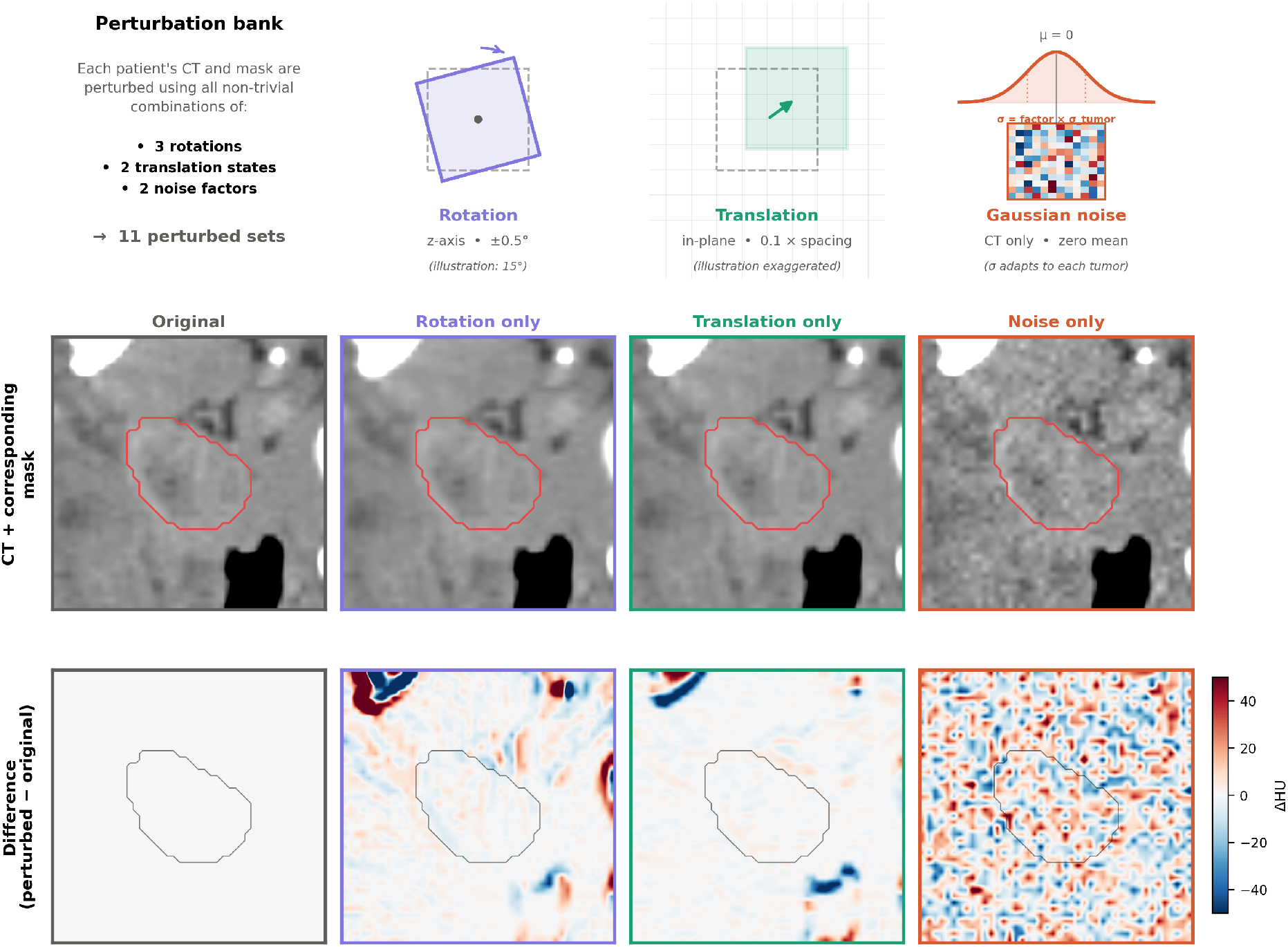
Overview of the perturbation framework used to generate perturbed CT image sets for radiomic repeatability analysis. The top row schematically illustrates the three perturbation components: small in-plane rotations around the z-axis, sub-voxel in-plane translations, and additive Gaussian noise applied to the CT image only. For each patient, combinations of these components were used to generate multiple perturbed image sets from the original CT image and tumor mask. The middle row shows representative axial slices of the original CT and three single-perturbation cases (rotation only, translation only, noise only) with the corresponding tumor contours overlaid in red. The bottom row presents voxel-wise intensity difference maps (*I*_perturbed_ − *I*_original_), revealing the spatial signature of each perturbation type: rotation produces symmetric intensity bands at tissue boundaries, translation produces faint directional gradients at sub-voxel scale, and Gaussian noise produces a stochastic pattern across the field of view.

**Figure 3:**
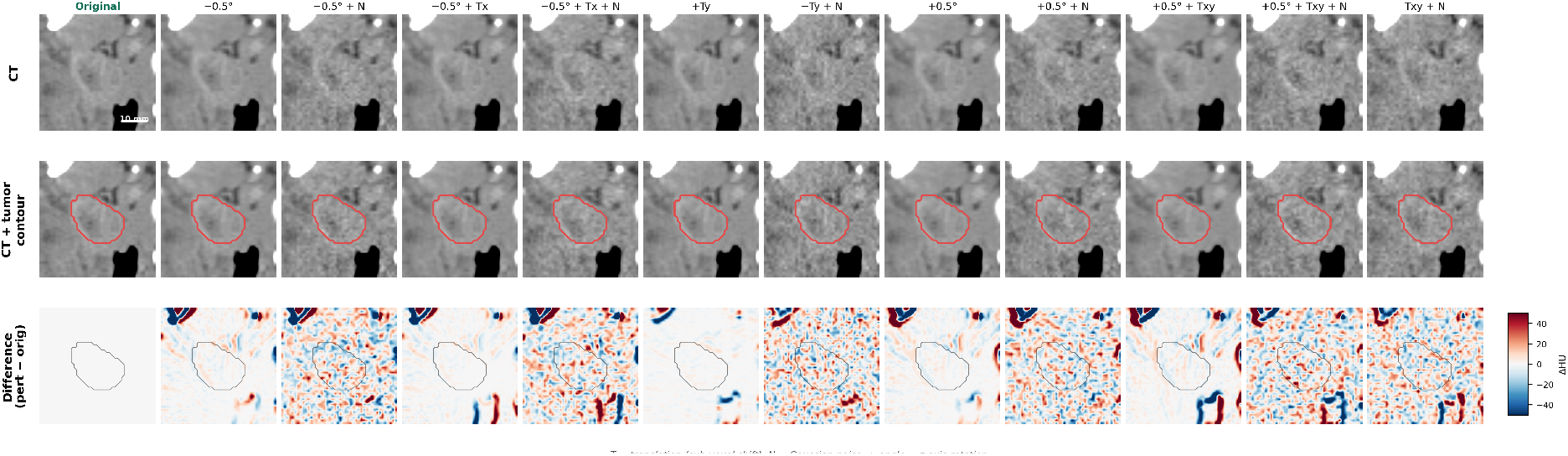
Visualization of the complete perturbation bank applied to a representative patient. The first row shows the original CT image and all 11 perturbed CT images generated by the perturbation framework. The second row shows the corresponding tumor contours, where rotations and translations are applied jointly to both the CT and the segmentation mask, while noise is applied only to the CT image. The third row presents voxel-wise intensity difference maps (*I*_perturbed_ − *I*_original_). Together, these panels illustrate that the perturbations introduce controlled low-level variability while preserving the overall anatomical structure and the spatial correspondence between CT images and tumor masks.

For each patient, multiple perturbed versions of the original CT image were generated. A total of 11 perturbed images were created in addition to the original image, resulting in 12 images per patient. The perturbation parameters were chosen following the protocols of Prior et al. [20] and the single-perturbation (SP) scenario of Bernatowicz et al. [19], both of which adopt small in-plane rotations (0.5°), sub-voxel translations (0.1× voxel spacing), and image-derived Gaussian noise as a minimal but representative perturbation set for radiomics repeatability assessment. The applied perturbations therefore consisted of small in-plane rotations, sub-voxel translations, and additive Gaussian noise. Specifically, rotation around the z-axis took values from {−0.5°, 0.0°, +0.5°}, translation was defined as a fraction of voxel spacing {0.0, 0.1} applied along fixed signed in-plane directions, and the addition of Gaussian noise was controlled by a binary factor {0, 1}. The full factorial combination of these parameters yielded 3 × 2 × 2 = 12 settings; excluding the no-perturbation case (which corresponds to the original image) resulted in 11 distinct perturbations per patient. Following the observation by Zwanenburg et al. [26] that noise addition shows only a marginal effect when combined with other perturbations, a separate noise-only configuration was not evaluated as an isolated case; instead, noise effects are assessed across six configurations spanning different rotation and translation contexts within the bank. The number of perturbations (11 per patient) was chosen to balance computational cost against directional coverage, extending the single-perturbation approach of Bernatowicz et al. [19] to a deterministic bank that spans all three rotation values, both translation magnitudes, both noise factors, and eight signed in-plane translation directions, thereby ensuring that feature stability is not biased toward any single shift direction. The complete list of the 11 applied perturbations is summarized in Table 1.

**Table 1:** Parameters of the 11 image perturbations applied to each patient. Rotations are around the z-axis (axial plane), translations are expressed as a fraction of voxel spacing, and noise factor of 1 corresponds to a Gaussian noise standard deviation equal to the standard deviation of CT intensities within the tumor mask.

| Perturbation ID | Rotation | Translation | Noise factor | Direction |
| --- | --- | --- | --- | --- |
| Perturbation 1 | $-0.5^\circ$ | none | 0 | – |
| Perturbation 2 | $-0.5^\circ$ | none | 1 | – |
| Perturbation 3 | $-0.5^\circ$ | $0.1 \times$ spacing | 0 | $+x$ |
| Perturbation 4 | $-0.5^\circ$ | $0.1 \times$ spacing | 1 | $-x$ |
| Perturbation 5 | $0^\circ$ | $0.1 \times$ spacing | 0 | $+y$ |
| Perturbation 6 | $0^\circ$ | $0.1 \times$ spacing | 1 | $-y$ |
| Perturbation 7 | $+0.5^\circ$ | none | 0 | – |
| Perturbation 8 | $+0.5^\circ$ | none | 1 | – |
| Perturbation 9 | $+0.5^\circ$ | $0.1 \times$ spacing | 0 | $+x, +y$ |
| Perturbation 10 | $+0.5^\circ$ | $0.1 \times$ spacing | 1 | $-x, +y$ |
| Perturbation 11 | $0^\circ$ | $0.1 \times$ spacing | 1 | $+x, -y$ |

Gaussian noise was applied to the CT image only, with zero mean. The standard deviation of the applied noise (*σ*_used_) was defined as:

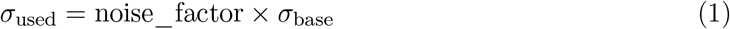

where *σ*_base_ represents the standard deviation of CT intensities within the tumor mask of each patient. This patient-adaptive scaling ensures that the noise magnitude is proportional to the intrinsic intensity heterogeneity of each tumor, allowing fair comparison of feature stability across tumors with widely varying levels of heterogeneity.

Rotation and translation transformations were applied jointly to each CT volume and its tumor mask, maintaining their geometric alignment. Grayscale CT volumes were resampled via linear interpolation, whereas binary segmentation masks underwent nearest-neighbor mapping to retain discrete label values. After each perturbation, all volumes were re-mapped onto the original CT geometry (matching size, spacing, origin, and direction), thereby ensuring voxel-wise correspondence by design. Inter-image registration was not required, given that the applied perturbation parameters were known a priori.

### 2.3 Voxelwise Radiomics Feature Extraction

Following the construction of the perturbation bank, voxelwise radiomics feature maps were extracted for every image set. In contrast to conventional segment-level (lesion-level) radiomics, where a single scalar value is computed per feature for the entire region of interest, voxelwise extraction computes a feature value *at every voxel* located inside the tumor mask [19, 20]. Each feature is therefore represented as a three-dimensional parametric map of the same geometry as the underlying CT volume, in which intratumoral spatial heterogeneity is preserved and can be quantitatively analyzed. This representation is essential for the subsequent voxel-level reproducibility analysis, since it allows perturbation-induced variability to be evaluated locally rather than after aggregation over the whole lesion. Feature extraction was implemented in Python using the open-source PyRadiomics library [27], which is fully compliant with the Image Biomarker Standardisation Initiative (IBSI) reference definitions [28]. For every patient, twelve image sets — one original and eleven perturbed versions — were processed independently using identical extractor settings, so that any differences observed between the resulting feature maps could be attributed solely to the imposed perturbations and not to differences in the extraction pipeline. The complete set of parameters is summarized in Table 2.

**Table 2:** Voxelwise radiomics feature extraction parameters. Settings follow the R1B25 protocol and are applied identically to the original and all eleven perturbed image sets of each patient.

| Category | Parameter | Value |
| --- | --- | --- |
| Protocol | Configuration label | R1B25 |
| Discretization | Bin width | 25 HU (fixed bin width) |
| Kernel | Kernel radius | 1 voxel ($3 \times 3 \times 3$ spherical, 27 voxels) |
| Kernel | Outside-mask init value | NaN |
| Feature classes | Enabled | firstorder, GLCM, GLRLM, GLSZM, GLDM, NGTDM |
| Feature classes | Disabled | Shape (no voxelwise definition) |
| Output | Map format | 3D NRRD |
| Image sets | Per patient | 1 original + 11 perturbed = 12 |
| Maps | Per image set | 93 |

The extraction protocol followed the so-called R1B25 configuration, which combines a kernel radius of one voxel with a fixed-bin-width gray-level discretization of 25 HU. With a kernel radius of one voxel and a spherical neighborhood definition, each voxel-level feature was computed over its local 3 × 3 × 3 (27-voxel) environment. The kernel was operated in *masked* mode, meaning that only voxels lying inside the tumor mask contributed to the local computation and voxels falling outside the mask were excluded from the neighborhood statistics. Voxels outside the mask in the output maps were assigned a not-a-number (NaN) value rather than zero, in order to avoid contaminating downstream statistics with artificial background values. A fixed bin width of 25 HU was used for gray-level discretization of the image intensities prior to texture matrix computation. This choice is consistent with established recommendations for CT-based radiomics, where a fixed bin width is preferred over a fixed bin count because it preserves the absolute physical meaning of the Hounsfield Unit scale across patients [29, 30, 28]. For each image set, the extractor returned one three-dimensional SimpleITK image per feature, which was written to disk in the NRRD format. 93 feature maps were produced per image set, yielding twelve sets of maps per patient. Figure 4 illustrates voxelwise feature maps for a representative patient across the original image set and 11 perturbed image sets, showing one representative feature from each radiomic feature family.

**Figure 4:**
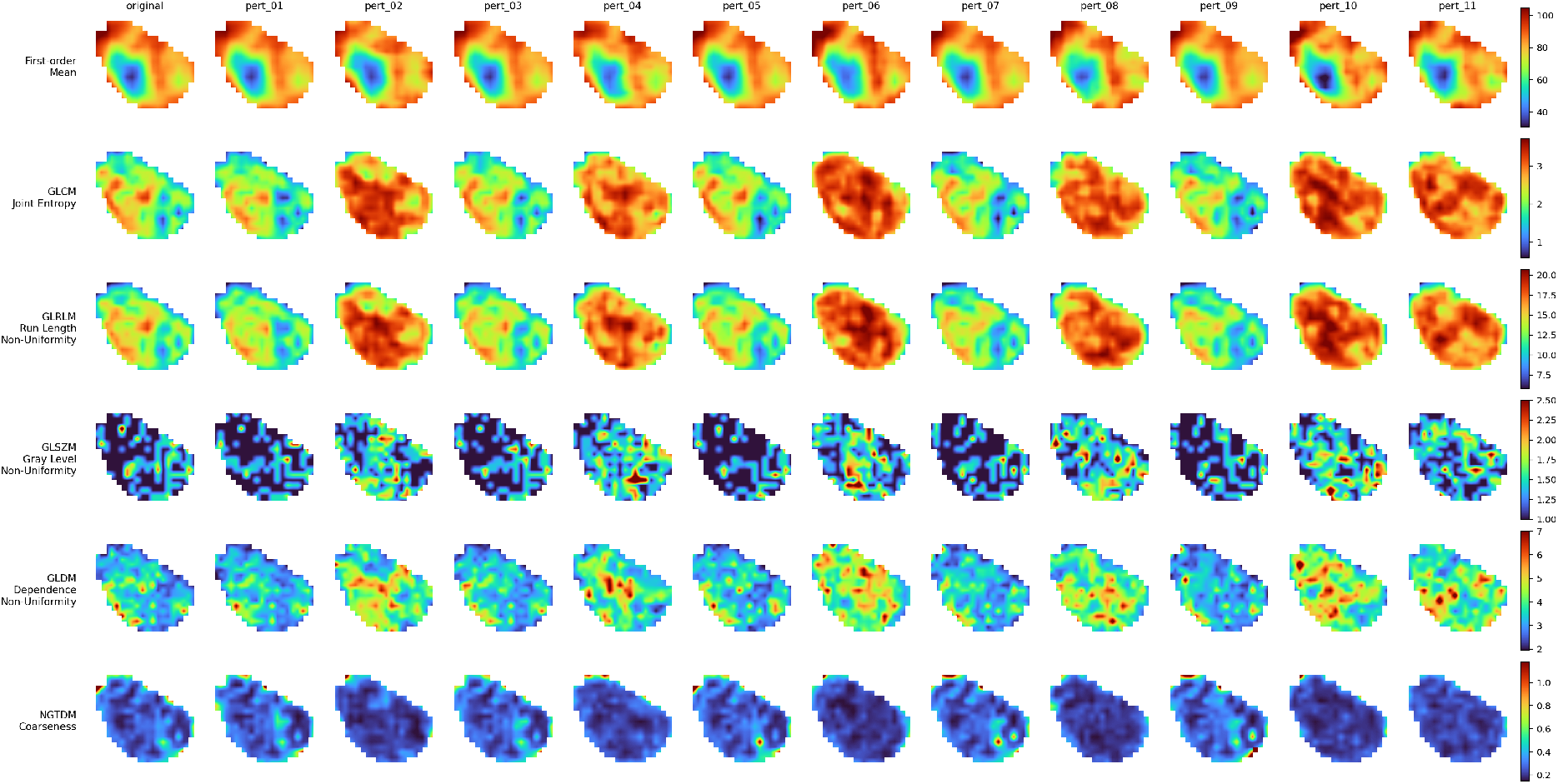
Voxelwise feature maps for a representative patient across the original image set and 11 perturbed image sets, showing one representative feature from each radiomic feature family: First-order (Mean), GLCM (Joint Entropy), GLRLM (Run Length Non-Uniformity), GLSZM (Size Zone Non-Uniformity), GLDM (Dependence Non-Uniformity), and NGTDM (Coarseness).

### 2.4 Assessment of Radiomic Feature Repeatability

Radiomic feature repeatability was quantified at the voxelwise level using the intraclass correlation coefficient (ICC), specifically the ICC(1,1) because the repeated measurements are exchangeable random realizations of the same perturbation process rather than systematically identifiable raters [31, 32]. For each patient, every voxelwise feature map was computed once on the original CT image and once on each of eleven independently perturbed versions of the same image, yielding *K* = 12 repeated voxel-wise measurements per feature; voxels with nonfinite values were treated as outside the region of interest, voxel intensities were rescaled to [0, 1] within each measurement using min–max normalization to ensure comparability across features, and the resulting vectors were trimmed to a common length to obtain a balanced design before computing ICC. To ensure a conservative estimation of feature stability, the lower bound of the 95% confidence interval of ICC (denoted as ICC_LCL) was used rather than the point estimate [33], since the LCL guarantees the minimum reliability that can be claimed at the 95% level and automatically penalizes estimates derived from small regions of interest. Based on ICC_LCL values, features were categorized into four groups:

- **Poor repeatability:** ICC_LCL *<* 0.50
- **Moderate repeatability:** 0.50 ≤ ICC_LCL *<* 0.75
- **Good repeatability:** 0.75 ≤ ICC_LCL *<* 0.90
- **Excellent repeatability:** ICC_LCL ≥ 0.90

This classification enabled the stratification of features into subsets with varying levels of robustness [16], which were subsequently used for habitat generation.

### 2.5 Computation of Habitats and Their Stability

Tumor habitats were generated using unsupervised clustering based on voxel-wise radiomic features. For each feature subset (All, Excellent, Good, Moderate, and Poor), a separate clustering process was performed. K-means clustering [34, 35] was applied to group voxels within the tumor into *H* = 3 distinct habitats, representing subregions with similar radiomic characteristics. The choice of *H* = 3 was based on prior analysis and consistency with established habitat imaging studies [19, 20]. The detailed habitat generation pipeline is described in our previous work [36]. In this study, the same framework was applied to generate habitat maps for both original and perturbed images. This process resulted in multiple habitat maps per patient, each corresponding to a different feature subset and perturbation instance.

Due to the unsupervised nature of clustering, habitat labels are inherently unordered, meaning that label assignments may differ across perturbations. To address this issue, an optimal matching strategy was applied to align habitat labels between original and perturbed maps. The Hungarian algorithm was used to determine the optimal one-to-one correspondence between habitats by maximizing spatial overlap [37, 38]. This step ensures that the same anatomical regions are compared across perturbations, enabling accurate computation of stability metrics.

For each patient, habitat stability was quantified using the Dice Similarity Coefficient (DSC) [39, 40] between habitat maps derived from original and perturbed images. For a given perturbation *k*, the DSC was first computed for each matched habitat after optimal matching. Each perturbation produces one DSC value by comparing the original and perturbed habitat maps.

The perturbation-level DSC was then defined as the average overlap across all *H* habitats:

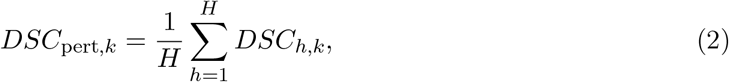

where

- *DSC*_*h,k*_ is the Dice Similarity Coefficient for habitat *h* under perturbation *k*,
- *H* is the total number of habitats (in this study, *H* = 3).

For each patient, the final DSC value was defined as the median across all *N* perturbations:

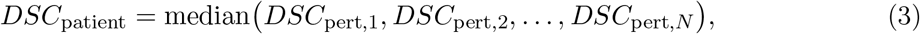

where *N* is the total number of perturbations applied to each patient.

In the discovery cohort, this habitat generation pipeline was applied across all five feature subsets (All, Excellent, Good, Moderate, Poor) defined by the cohort-specific ICC_LCL classification. In the external validation cohort, the same pipeline was applied using the *fixed* feature assignments derived from the discovery cohort, without any re-fitting of the stability classification, so that the validation cohort served exclusively as a test of generalizability.

### 2.6 Statistical Analysis

Group-wise differences in habitat stability were probed through Wilcoxon rank-sum tests applied pairwise across the feature subsets. The customary *p <* 0.05 threshold defined statistical significance, yet every pairwise outcome exceeded this criterion, falling below *p <* 0.001.

### 2.7 External Validation Strategy

The generalizability of the proposed stability framework was assessed in the independent validation cohort using three complementary perspectives. First, at the aggregate level, the overall distribution of features across the four stability categories (poor, moderate, good, excellent) was compared between the discovery and validation cohorts to evaluate whether the proportion of features in each category was preserved across cohorts. Second, at the feature level, the agreement between the two cohorts was quantified by treating each feature’s category in the discovery cohort as the reference label and comparing it to its category in the validation cohort. Overall agreement, unweighted Cohen’s *κ*, and quadratic-weighted Cohen’s *κ* were computed across all 93 features, and per-class one-versus-rest *κ* values were additionally reported for each of the four categories. The quadratic-weighted formulation was included to account for the ordered nature of the stability categories, in which misclassifications to adjacent categories are less severe than misclassifications across the full scale. Third, at the functional level, the discovery-derived stability classes (excellent, good, moderate, poor) were used as fixed feature subsets to generate habitat maps in the validation cohort, and the resulting per-patient DSC distributions were compared across subsets using the same pairwise Wilcoxon rank-sum tests described in Section 2.6. This three-level design enabled us to evaluate whether the stability classification reproduces in distribution, whether it reproduces at the level of individual features, and whether it reproduces in its downstream functional consequence on habitat stability.

## 3 Results

### 3.1 Radiomic Feature Repeatability

The distribution of radiomic feature repeatability in the discovery cohort is presented in Figure 5. Overall, a large proportion of features exhibited low repeatability, with 48.4% classified as poor, 21.5% as moderate, 23.7% as good, and only 6.5% as excellent. This indicates that nearly half of the extracted radiomic features are sensitive to image perturbations. When stratified by feature class, repeatability varied across feature groups. First-order features showed relatively higher stability, with a greater proportion of features in the good and excellent categories compared to other feature classes. In contrast, texture-based features demonstrated lower repeatability. In particular, GLSZM and GLRLM features exhibited a higher proportion of poorly repeatable features, with 75.0% and 62.5% classified as poor, respectively. Similarly, GLDM features showed a predominance of poor repeatability (50.0%), while GLCM features were more evenly distributed across repeatability categories. NGTDM features, although limited in number, showed relatively higher stability, with 60.0% classified as good. These results demonstrate that feature repeatability is not uniform across feature classes and that higher-order texture features are generally more sensitive to perturbations.

**Figure 5:**
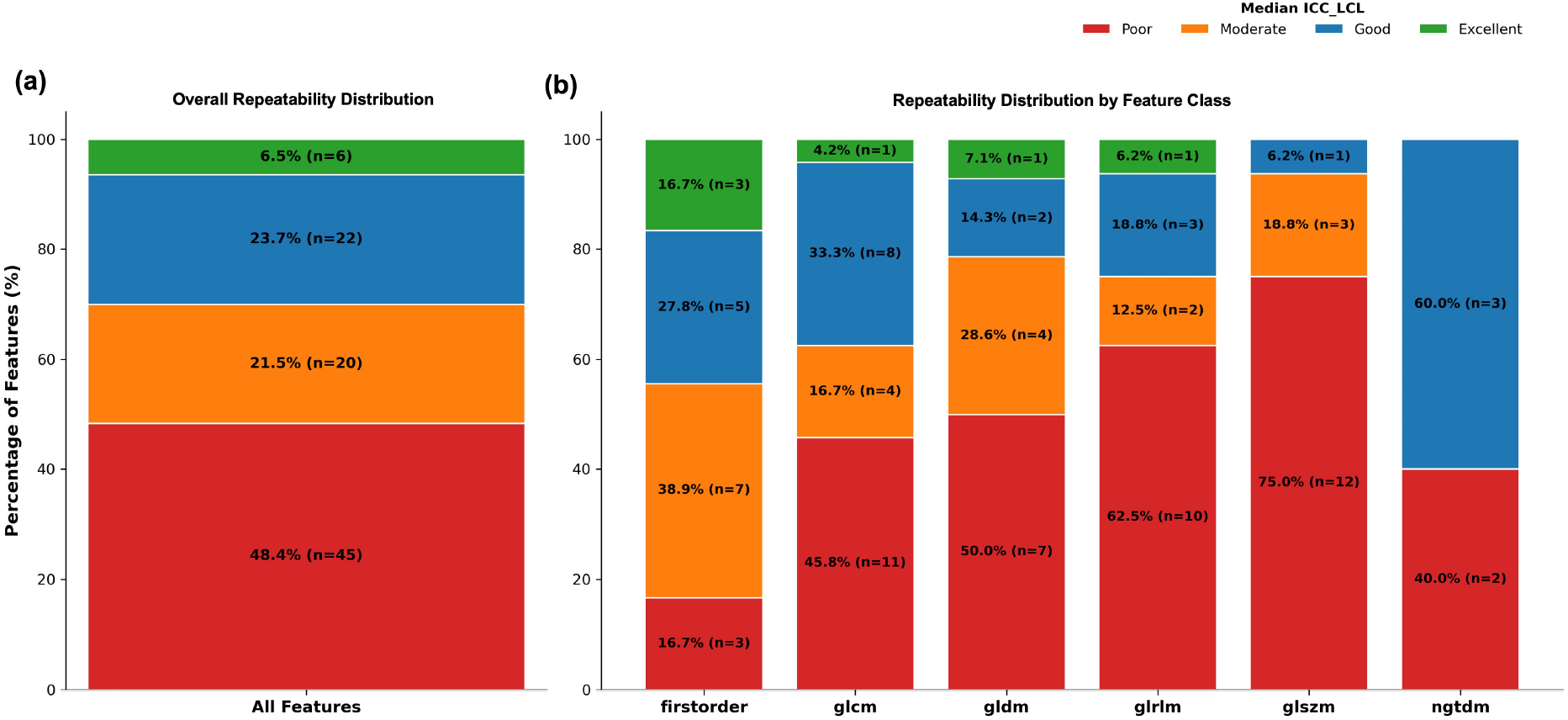
Radiomic feature distributions in the discovery cohort based on ICC_LCL(1,1) after 11 different image perturbations per image. (a) Repeatability across all radiomic features. (b) Feature stability grouped by radiomic class (first-order, GLCM, GLDM, GLRLM, GLSZM, NGTDM).

### 3.2 Habitat Stability Across Feature Subsets

The impact of feature repeatability on habitat stability is shown in Figure 6a. Habitat stability was quantified using the Dice Similarity Coefficient (DSC) between habitat maps derived from original and perturbed images. Significant differences in habitat stability were observed across feature subsets (all pairwise comparisons: *p <* 0.001). Habitats generated using features with excellent repeatability showed the highest stability, with median DSC values close to 0.9 and a relatively narrow interquartile range. Features with good repeatability also produced stable habitat maps, although with slightly greater variability compared to the excellent group. In contrast, habitats generated from moderately repeatable features exhibited lower stability, with a broader distribution of DSC values. The lowest stability was observed for habitats derived from poorly repeatable features, with median DSC values around 0.4 and increased variability across patients. Habitats generated using all features showed intermediate stability, with median DSC values around 0.5, suggesting that the inclusion of less repeatable features may reduce the overall consistency of habitat maps. Overall, these results show a clear trend of decreasing habitat stability with decreasing feature repeatability.

**Figure 6:**
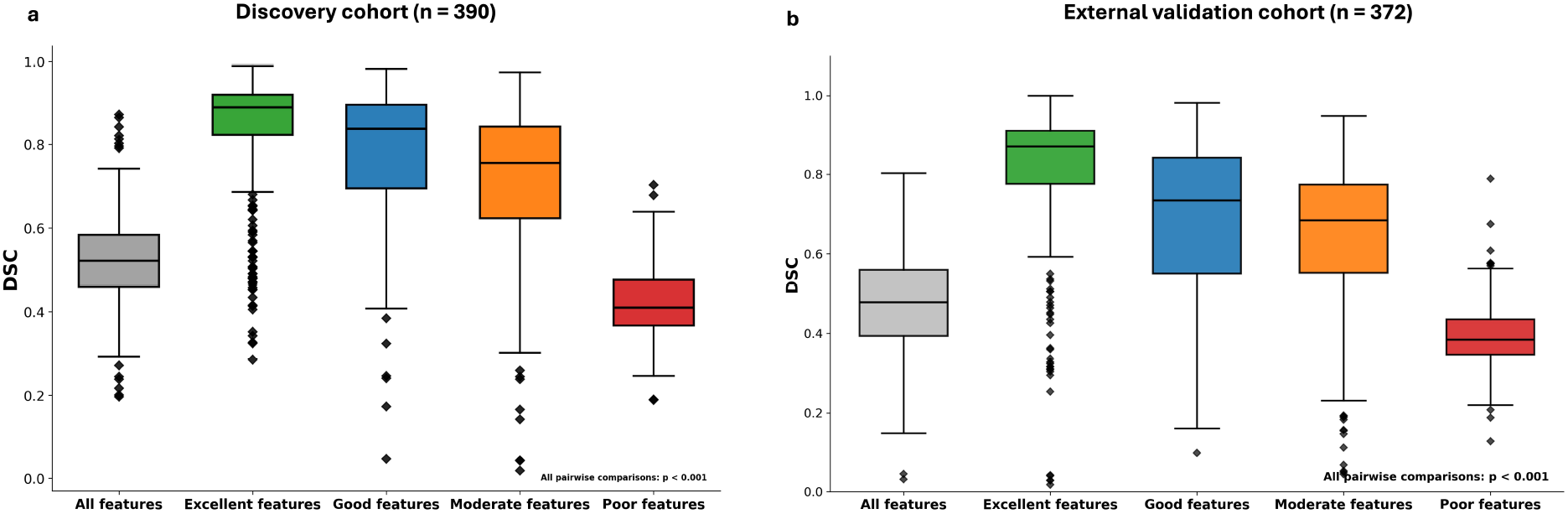
Per-group habitat stability across radiomic feature subsets defined by ICC_LCL thresholds in (a) the discovery cohort (*n* = 390) and (b) the external validation cohort (*n* = 372). Five subsets were considered: *All features* (no filtering), *Excellent* (ICC_LCL ∈ [0.90, 1]), *Good* (ICC_LCL ∈ [0.75, 0.90)), *Moderate* (ICC_LCL ∈ [0.50, 0.75)), and *Poor* (ICC_LCL *<* 0.50). For every patient, the Dice coefficient was computed between habitat maps generated from the original CT volume and those from each perturbed counterpart, and the per-patient median across perturbations served as the stability score. The monotonic relationship between feature repeatability and habitat stability observed in the discovery cohort is fully preserved in the external validation cohort, with the same ordering of median DSC values (excellent *>* good *>* moderate *>* poor). Boxplots summarize the within-subset distribution; isolated points indicate outliers. Wilcoxon testing showed that all pairwise contrasts attained *p <* 0.001 in both cohorts.

### 3.3 External Validation in the Institutional Cohort

The generalizability of the proposed stability framework was evaluated in the independent institutional cohort of 372 oropharyngeal cancer patients, following the three-level design described in Section 2.7.

At the aggregate level, the overall distribution of features across the four stability categories was largely preserved between the two cohorts (Figure 7b). The proportion of features classified as poorly repeatable was nearly identical in the two cohorts (48.4% in discovery vs. 49.5% in validation), while a modest downward shift was observed in the higher categories, with the validation cohort showing a slightly larger fraction of moderately repeatable features (33.3% vs. 21.5%) and a smaller fraction of good (15.1% vs. 23.7%) and excellent (2.2% vs. 6.5%) features. The relative ranking of feature classes was preserved, with first-order features remaining the most repeatable and GLSZM features remaining the least repeatable in both cohorts.

**Figure 7:**
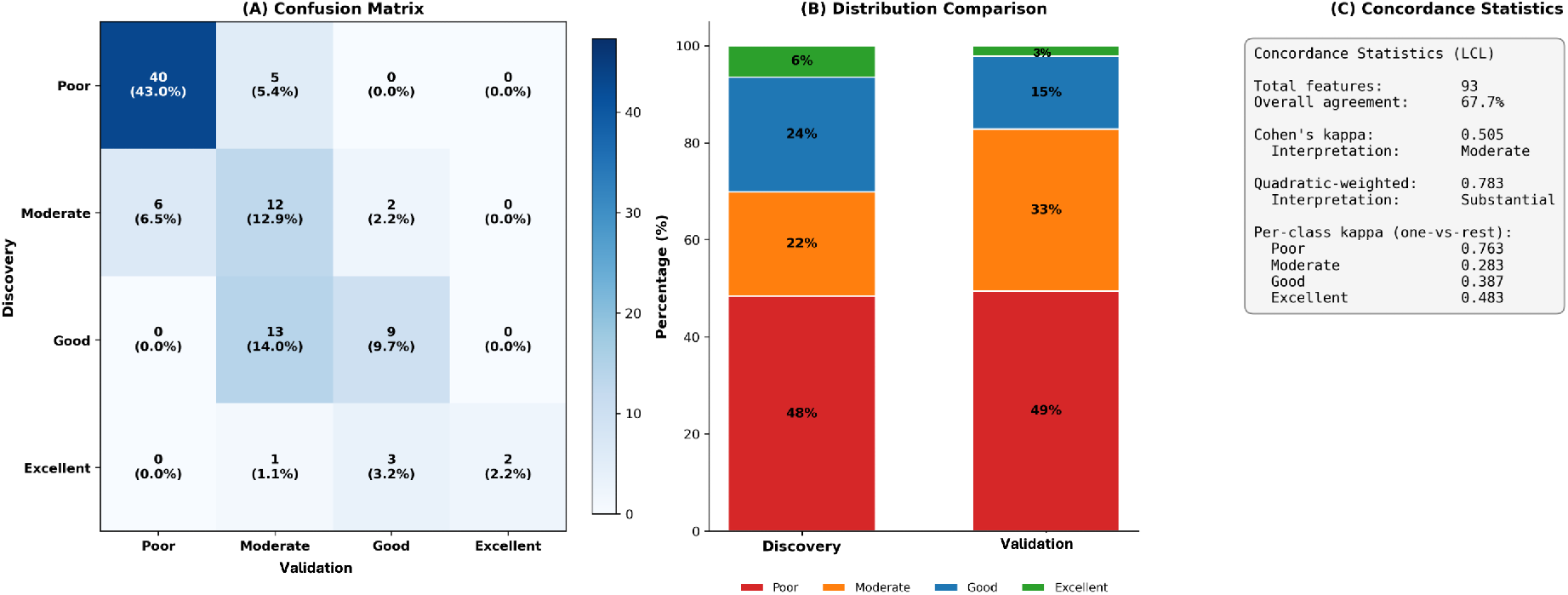
Concordance of the radiomic feature stability classification between the discovery and external validation cohorts. (a) Confusion matrix showing the cross-tabulation of stability categories assigned to each of the 93 features in the two cohorts, with diagonal entries corresponding to features classified into the same category in both cohorts; all off-diagonal entries are confined to adjacent categories, and no feature shifts by more than two stability classes. (b) Stacked distribution of features across the four stability categories in each cohort, illustrating the preserved overall balance between repeatability categories. (c) Concordance statistics, including the overall percentage of features assigned to the same category, the unweighted and quadratic-weighted Cohen’s *κ* with their interpretation, and the per-class one-versus-rest *κ* values. Classification was based on ICC_LCL using the same thresholds as in Figure 5.

At the feature level, the agreement between cohorts was substantial. The overall percentage of features assigned to the same stability category in both cohorts was 67.7%, with an unweighted Cohen’s *κ* of 0.505 (moderate agreement) and a quadratic-weighted *κ* of 0.783 (substantial agreement). Per-class one-versus-rest analysis showed that the poor category had the strongest concordance (*κ* = 0.76), followed by the excellent (*κ* = 0.48), good (*κ* = 0.39), and moderate (*κ* = 0.28) categories. Importantly, no feature shifted by more than two stability classes between the two cohorts: all disagreements were confined to adjacent categories (Figure 7a). The discrepancy between the unweighted and quadratic-weighted *κ* therefore reflects local boundary effects between adjacent stability categories rather than systematic misclassification across the full scale.

At the functional level, habitats generated in the validation cohort using the discovery-derived stability classes reproduced the same monotonic relationship between feature repeatability and habitat stability as the discovery cohort (Figure 6b). The median DSC was 0.87 for habitats generated from excellent features, 0.73 for good features, 0.69 for moderate features, and 0.39 for poorly repeatable features, while habitats generated from all features yielded an intermediate median DSC of 0.48. All pairwise comparisons between feature subsets remained statistically significant (*p <* 0.001). The absolute DSC values were slightly lower in the validation cohort than in the discovery cohort, consistent with the broader distribution of imaging protocols and scanners in the institutional setting, but the ordering of feature subsets was fully preserved.

## 4 Discussion

In this study, we evaluated how the repeatability of voxel-wise radiomic features affects the spatial stability of tumor habitats in HNC using a perturbation-based framework. Our analysis on 390 oropharyngeal cancer patients showed that nearly half of the extracted voxel-wise features (48.4%) were classified as poorly repeatable, while only 6.5% reached the excellent category. More importantly, this feature-level variability propagated directly to the habitat level: habitats generated from excellent features achieved a median Dice similarity coefficient (DSC) close to 0.9, whereas habitats based on poorly repeatable features dropped to a median DSC near 0.4. Habitats built from the full feature set fell in between (median DSC ≈ 0.5), indicating that including unstable features dilutes the spatial consistency of the resulting maps.

Our findings are consistent with previous voxel-wise repeatability analyses showing that radiomic feature stability varies across feature types and tumor sites. Bernatowicz et al. [19] reported that, in lung lesions from the RIDER cohort, only a limited subset of voxel-wise GLCM features, including joint energy, joint entropy, and sum entropy, reached the high-repeatability range. Prior et al. [20] evaluated 2,436 lung and liver lesions using perturbation-based analysis and stratified voxel-wise features into poor, moderate, good, and excellent repeatability categories based on ICC_LCL. For habitat-stability analysis, however, features were primarily grouped as precise versus non-precise using an ICC_LCL threshold of ≥ 0.50. In our study, 51.6% of features achieved at least moderate repeatability using the same ICC_LCL threshold. More importantly, the four repeatability categories were carried forward directly into the habitat-stability analysis, allowing habitat stability to be evaluated across multiple levels of feature robustness rather than a binary grouping. This stratified design showed a clear monotonic relationship between feature repeatability and habitat stability. Habitats generated using the complete feature set showed lower stability than habitats generated from highly repeatable features, indicating that repeatability-based feature selection can improve habitat reproducibility without modifying the clustering framework itself. The same hierarchical pattern was reproduced in the external validation cohort, supporting the reproducibility of the observed relationship across independent cohorts.

The differences observed between feature classes can be interpreted in light of how each feature is computed. First-order features summarize the intensity distribution within a local neighborhood and are mathematically less sensitive to small spatial perturbations, which explains the higher proportion of features in the good and excellent categories. In contrast, GLSZM and GLRLM features depend on connected regions or runs of voxels with similar intensity levels, and even small changes in voxel position or noise can break or merge these regions, leading to large changes in the resulting feature values. This is consistent with previous reports that texture features, particularly those based on size zones and run lengths, are more sensitive to image acquisition and preprocessing variability [16, 14]. Importantly, our analysis adds a new perspective by showing that this sensitivity is not only a feature-level concern: when these unstable features are used as input for habitat clustering, the resulting tumor subregions also become spatially unstable.

From a methodological perspective, our findings reinforce the value of perturbation-based frameworks for radiomics quality control. Test–retest CT data are rarely available in clinical practice, and existing public datasets cover only a limited number of anatomical sites [17]. Image perturbation, as originally proposed by Zwanenburg et al. [26], offers a practical alternative that can be applied to any single-acquisition cohort. Our deterministic perturbation bank, which combines small in-plane rotations, sub-voxel translations, and tumor-adaptive Gaussian noise, ensures directional coverage of translations and a controlled noise scale adapted to each tumor. This design provides a reproducible basis for stability analysis and can be applied to other cancer types or imaging modalities with minimal modification. Our results also support the recommendation, already made in earlier studies [19, 20], that radiomics workflows should incorporate a feature-stability filtering step before downstream analysis; in the specific context of habitat imaging, this filtering directly improves the spatial reproducibility of the resulting maps.

Importantly, all three observations made in the discovery cohort generalized to the independent institutional cohort. The overall distribution of features across stability categories was largely preserved between the two cohorts, and the relative ranking of feature classes (first-order being the most repeatable and GLSZM the least) was reproduced. At the level of individual features, the agreement was substantial (quadratic-weighted *κ* = 0.78), and no feature shifted by more than two stability classes, indicating that the classification is robust to scanner and protocol differences and that disagreements are limited to local boundary effects between adjacent categories. Finally, when the discovery-derived stability classes were used as fixed feature subsets in the validation cohort, the monotonic relationship between feature repeatability and habitat stability was fully preserved, with the same ordering of median DSC values (excellent *>* good *>* moderate *>* poor). The absolute DSC values were slightly lower in the validation cohort, which is consistent with the broader range of CT acquisition protocols in an institutional setting, but the ordering and the statistical separation between subsets were maintained. Together, these observations indicate that the proposed stability classification generalizes beyond the discovery cohort and provides a transferable basis for selecting features used in HNC habitat imaging.

This study has some limitations. The proposed stability framework quantifies the spatial reproducibility of habitat maps but does not assess their biological accuracy, as ground-truth data such as whole-mount histopathological mapping or co-registered multi-modal imaging were not available. Validating habitat accuracy through histopathological correlation is therefore an important next step that we plan to address in future work.

## 5 Conclusion

This study shows that radiomic feature repeatability has a direct and substantial impact on habitat stability in head and neck cancer. Habitat consistency scales monotonically with the underlying feature repeatability, with median DSC values ranging from approximately 0.9 for excellent features down to 0.4 for poorly repeatable ones, and this monotonic relationship was reproduced in an independent institutional cohort of 372 oropharyngeal cancer patients. These findings indicate that selecting features with higher repeatability can substantially improve the spatial reproducibility of habitat maps without modifying the clustering pipeline, and they support the integration of perturbation-based stability analysis as a standard step in habitat imaging workflows for HNC.

## Data Availability

All data produced in the present study are available upon reasonable request to the authors.

## Acknowledgments

This work has been supported in part by the Collaborative Data Services Core (CDSC) and Quantitative Imaging Core (QIC) Core at the H. Lee Moffitt Cancer Center & Research Institute, an NCI designated Comprehensive Cancer Center (P30 CA076292).

